# gene.iobio: an interactive web tool for versatile, clinically-driven variant interrogation and prioritization

**DOI:** 10.1101/2020.11.05.20224865

**Authors:** Tonya Di Sera, Matt Velinder, Alistair Ward, Yi Qiao, Stephanie Georges, Chase Miller, Anders Pitman, Will Richards, Aditya Ekawade, David Viskochil, John C Carey, Laura Pace, Jim Bale, Stacey L Clardy, Ashley Andrews, Lorenzo Botto, Gabor Marth

## Abstract

With increasing utilization of comprehensive genomic data to guide clinical care, anticipated to become the standard of care in many clinical settings, the practice of diagnostic medicine is undergoing a notable shift. However, the move from single-gene or panel-based genetic testing to exome and genome sequencing has not been matched by the development of tools to enable diagnosticians to interpret increasingly complex genomic findings. A new paradigm has emerged, where genome-based tests are often evaluated by a large multi-disciplinary collaborative team, typically including a diagnostic pathologist, a bioinformatician, a genetic counselor, and often a subspeciality clinician. This team-based approach calls for new computational tools to allow every member of the clinical care provider team, at varying levels of genetic knowledge and diagnostic expertise, to quickly and easily analyze and interpret complex genomic data. Here, we present *gene.iobio*, a real-time, intuitive and interactive web application for clinically-driven variant interrogation and prioritization. We show *gene.iobio* is a novel and effective approach that significantly improves upon and reimagines existing methods. In a radical departure from existing methods that present variants and genomic data in text and table formats, *gene.iobio* provides an interactive, intuitive and visually-driven analysis environment. We demonstrate that adoption of *gene.iobio* in clinical and research settings empowers clinical care providers to interact directly with patient genomic data both for establishing clinical diagnoses and informing patient care, using sophisticated genomic analyses that previously were only accessible via complex command line tools.

## Introduction

It is becoming increasingly common for clinical care providers to incorporate genetic information into a patient’s clinical diagnosis and subsequent care. This major transition in care relies on a number of factors, including but not limited to: a patient’s access to genetic sequencing; genetics education of providers and an understanding of how genetic variants can impact patient care; time constraints on clinicians and clinical groups to adopt additional considerations and workloads into their patient care; and clinicians’ ability to confidently analyze and interpret genetic findings. Given the wide scope of this clinical transition, we developed *gene.iobio* to address the challenge clinicians face in analyzing and interpreting genomic findings.

Typical exome or genome sequencing studies produce vast amounts of data that are stored on cloud or institutional hardware. Raw sequencing reads pass through a number of complex processing tasks to generate variant calls. For a typical trio exome sequencing study (proband, mother and father), it is expected that well over 50,000 variants will be identified. This number reaches multiple millions when the entire genome is analyzed. These variants need to be prioritized and evaluated based on whether they can reasonably contribute to the patient’s phenotype. Given the number of variants, this is a daunting task that must consider numerous factors such as: the Mendelian mode of inheritance and segregation of the given allele in the family, population allele frequency, predicted impact and biological consequence, known gene:disease associations and in-silico predictions of pathogenicity. Command line, UNIX-based, variant prioritization tools have been developed to consider these factors^1–4^. However, these tools are often difficult to download, install, and run - making it unreasonable for clinician care providers to perform these analyses. As a result, the current paradigm relies on a team of experts across multiple disciplines to deliver optimal and expedient genomically-informed care. This typically involves bioinformaticians, clinical geneticists, molecular pathologists and subspecialty clinicians - all of whom need to be in near-constant communication and have in-depth discussions about candidate variants before reaching a clinical diagnosis based on genetic information. This level of logistics and organization becomes even more challenging in sequencing studies that demand rapid turnover and timely diagnoses, as clinical interventions informed by genetic findings can significantly improve patient outcomes and prognoses^5^.

Broadly speaking, our approach to these challenges has been to reimagine the current paradigm and bring clinical knowledge closer to genomic data and variant interpretation. This approach has led to the development of an expanding suite of intuitive, visually-driven, web-based bioinformatics tools; the iobio^6^ software suite. Our other iobio tools provide rapid quality review of BAM/CRAM files^7^ (http://bam.iobio.io), and VCF files (http://vcf.iobio.io), and for generating lists of genes associated with given genetic disorders and phenotypes^8^ (http://genepanel.iobio.io). *Gene.iobio* expands our iobio approach into variant interrogation and prioritization. Few tools have attempted similar visual web-based approaches. The vast majority of these tools (Emedgene, Alamut, Fabric, Varsome Premium, QIAGEN Ingenuity, Genuity Science and others) have been commercialized with significant licensing costs - limiting access to academic research groups and underserved clinics and their patients. The only free-to-use academic option to our knowledge is VCF/Plotein^9^, which attempts to visualize variants and their context within publicly-available variant databases as well as their pathogenicity scores. However, the VCF/Plotein tool lacks significant functionality required for comprehensive variant prioritization in clinical settings. *Gene.iobio* addresses numerous unsolved clinical genetics challenges and provides a comprehensive, up-to-date, disease and phenotype-informed, variant review platform for clinicians and clinical teams. Even before a dedicated publication, the tool has become extremely popular and highly accessed, with >1,000 distinct monthly users, each performing an average of 3 analysis sessions 15 minutes or longer and involving >15 user interactions.

## Methods

### File input/output

G*ene.iobio* accepts file-format compliant indexed BAM/CRAM^10^ and indexed (unannotated or annotated) VCF^11^ files. Files can be provided via a publicly accessible URL, secure private URLs (via access tokens or VPN), and/or through the user’s local machine. Importantly, these files can be in distinct locations (e.g. BAM/CRAM files on the user’s local machine and the VCF via a URL). Regardless of file locations, *gene.iobio* streams the relevant portions of data, including VCF variant data and BAM/CRAM sequencing data, and displays them in a visual interface for the user to analyze. *Gene.iobio* allows a user to save results as a VCF or a comma-separated values file through an “Export Variants” option. This exported file includes all relevant annotations and reviews made by the user for loading back into *gene.iobio* using the “Import Variants” option, allowing users to recall previously saved analyses at any time and across browser sessions. Additionally, *gene.iobio*, can save analyses back to Mosaic, a commercial and collaborative genomic data platform, developed by Frameshift Genomics (https://frameshift.io/).

### System architecture

*Gene.iobio* is a Javascript application that interfaces with our cloud-based iobio backend services (https://github.com/iobio/iobio-gru-backend). This architecture delineates application and data processing logic. *Gene.iobio* controls user visualizations and interactions in the browser and the coordination between various visual components, as well as interfacing with the iobio backend. The iobio backend services are functionally equivalent to command line bioinformatics tools, wrapped as web services. The iobio backend performs region-based bioinformatics analyses (see below) on source files (BAM/CRAM and VCF) and transforms these data into formats that are interpretable to web applications such as *gene.iobio*. These region-based analyses analyze only the streamed data, as discrete and manageable chunks, allowing the outputs to automatically update *gene.iobio* visualizations in real-time. *Gene.iobio* interfaces with iobio backend services asynchronously through secure HTTPS requests.

#### Variant annotation

Variant annotation is performed by iobio backend services in a region-specific manner, with the data streamed back to *gene.iobio*. This variant annotation service includes: *tabix*^*12*^ (for region-based querying of indexed VCF files), *vt*^*13*^ (for sample subsetting, variant decomposing, normalizing and transforming), *VEP*^*14*^ (for transcript-aware annotation of variants with functional consequence, impact, ClinVar^15^ significance, REVEL^16^ score, HGVS^17^, and dbSNP^18^ ID), and *bcftools* (for determining variant population allele frequency in gnomAD) (https://github.com/samtools/bcftools).

#### Sequencing data coverage and alignment

*Gene.iobio* displays sequencing data coverage visualizations based on the data returned from iobio backend services. This coverage-based iobio backend service utilizes *samtools*^*10*^ for region-based queries of CRAM/BAM alignment files and to determine coverage across a gene or a given region such as an exon. *Gene.iobio* visually summarizes coverage in these regions including the min, max, median and mean.

#### Variant calling

On-demand variant calling is performed by a backend service that includes *samtools*^*10*^ for region-based queries of CRAM/BAM alignment files and Freebayes^19^ for calling variants. Called variants are annotated in the same way as described in the Variant annotation section above.

#### IGV integration

*Gene.iobio* directly integrates a web-based JavaScript version of the Integrated Genome Viewer (IGV)^20^, called *igv.js* (https://github.com/igvteam/igv.js/).

#### Gene:disease association

*Gene.iobio* provides a controlled gene, phenotype and disorder vocabulary to help guide variant prioritization and ensure correct names have been loaded. *Gene.iobio* uses GENCODE^21^ and RefSeq^22^ gene names in an input text box with typeahead and autocomplete functionality. *Gene.iobio* also integrates Phenolyzer^23^, which allows the user to enter a phenotype term and automatically generate a list of genes associated with that phenotype. *Gene.iobio* retrieves up-to-date gene:disease association data from OMIM^24^ via their web API. PubMed articles associated with a particular gene are retrieved using the web API, NCBI E-utils.

### Language and codebase

*Gene.iobio* is a large and complex codebase with over 30,000 lines of code, and is available in the public GitHub repository at https://github.com/iobio/gene.iobio.vue. *Gene.iobio* uses the *Vue.js* Javascript framework that supports reusable components that are able to plug and play in different aspects of the application and more broadly within our suite of iobio applications. All interactive data visualizations are built using *D3*^*25*^, allowing for custom genomic visual components that respond in real-time to new data as it is streamed from iobio backend services.

#### Backend services

The *iobio backend* is written in server-side JavaScript running on Node.js. The source is available in the public GitHub repository (https://github.com/iobio/iobio-gru-backend). The iobio backend (gru) is responsible for remote invocation of command line bioinformatics pipelines. These pipelines are bash scripts that read from standard input and pipe through various bioinformatics applications and write data to standard output. Bash scripts are wrapped as web services using Node.js. Pipeline requests are made using remote procedure calls (RPCs), using simple Hypertext Transfer Protocol Secure (HTTPS) requests. Many of the requests are resource-intensive, therefore the gru iobio backend is designed to be horizontally scaled by load-balancing across as many compute nodes as necessary.

To serve local files, *gene.iobio* utilizes fibridge, a generic service that provides a way to proxy HTTP connections to a WebSocket source. The local file proxy greatly simplifies the code base, so that local files and remotely-service files are read from the iobio backend via an HTTP request. When the *gene.iobio* client application opens a local file it opens a WebSocket channel to the fibridge server. The fibridge server then provides a URL for the file.

The iobio backend heavily leverages Singularity containers, which allows each bioinformatics tool to be self-contained and isolated from the other tools as well as making them portable for other sites. This is particularly helpful for complex or legacy tools which may be written in languages like Perl, and for tools which have many dependencies. gru is loaded on AWS EC2 instances, providing a scalable, fault tolerant compute environment. The iobio backend is also installed in the University of Utah’s High Performance Computing Center’s protected environment.

### External resources and databases

*Gene.iobio* integrates numerous public datasets to present up-to-date gene and variant annotations. These external resources and databases are kept up-to-date using iobio backend services built around the individual data type. For instance, the ClinVar resource is maintained with a backend service that retrieves the latest ClinVar VCF on a weekly basis. ClinVar VCF variants are matched to user-provided variants during *gene.iobio* analyses. Similarly, the gnomAD resource is regularly updated and variants are matched during analyses. GnomAD population allele frequencies, as well as heterozygous and homozygous alternate allele counts, are provided within *gene.iobio*. Gene function summaries are retrieved via the NCBI E-utilities^26^ REST API. The iobio backend also maintains FASTQ files for genome reference builds including GRCh37 and GRCh38. The phlyoP^27^ conservation scores and multiple species sequence alignment visualizations in *gene.iobio* rely on UCSC^28^ genome tracks to display multiple organism sequence alignments surrounding the given variant. Gene names are maintained via GENCODE and RefSeq resources. Numerous other external links are provided at the gene- and variant-specific level, including MARRVEL^29^, VarSome^30^, OMIM^31^, DECIPHER^32^, GeneCards^33^, GTEx^34^ and others.

### Development

*Gene.iobio* has been developed using best practices in software development, versioning and testing. We maintain a dynamic codebase with multiple developers contributing to the project. The *gene.iobio* code base is maintained in GitHub. This allows changes to be merged into the current version, pull requests initiated and versions to be tracked. We have also developed a testing environment that allows us to deploy specific versions directly to the web via a unique URL. These test builds are automatically generated each time a pull request is submitted to the GitHub repository. This enables developers to quickly share their proposed changes with the rest of the team, making collaboration more efficient. We actively maintain the *gene.iobio* codebase, regularly making improvements, adding new features and deploying them to the public version on a regular release schedule.

### Deployment, usage and availability

*Gene.iobio* is publicly available and free-to-use for academic purposes at http://gene.iobio.io/. Commercial use is licensed through Frameshift Genomics (https://frameshift.io/). The University of Utah and the Utah Center for Genetic Discovery maintain an institutional version of *gene.iobio* for use by our clinical teams and genetics researchers. *Gene.iobio* has been developed and optimized for the Chrome browser, with additional support for the Firefox and Safari browsers.

*Gene.iobio* instances have been deployed at the University of Utah, Nebula Genomics^35^ (https://nebula.org/), MyGene2^36^ (https://mygene2.org/MyGene2/) and for an educational exhibit at the Natural History Museum of Utah (https://learngene.iobio.io/). From our web analytics, *gene.iobio* has more than 4,000 distinct users, with an average of 1,000 distinct users per month and more than 20,000 pageviews per month. The typical user performs multiple analysis sessions per month and spends more than 15 minutes per analysis session.

## Results

*Gene.iobio* is a real-time, intuitive and interactive platform for performing sophisticated gene and variant level review. *Gene.iobio* does not require time-consuming data uploads and can be used for real-time analysis of both exome and genome sequencing data. *Gene.iobio* can be used in singleton sequencing studies, but is most powerful in family studies where parents and additional siblings have been sequenced. For all variants in user-provided genes, *gene.iobio* determines allele segregation and visually displays the mode of inheritance, including the evidence for reference and alternate alleles. This allows users to enter suspected genes, given the clinical phenotype or from disease:gene association tools such as *genepanel.iobio*^*8*^, *Phenomizer*^*37*^ or *PanelApp*^*38*^ as well as genes prioritized by upstream variant prioritization tools such as slivar^4^, GEMINI^1^, ANNOVAR^2^ and others. We find an especially useful approach is to use *genepanel.iobio* to generate a comprehensive list of disease-associated genes and enter this gene list into *gene.iobio* for variant review. *Gene.iobio* provides visual summaries of pertinent variant annotations such as biological impact, gnomAD population frequency^39^, ClinVar assertion^15^, REVEL score^16^ and evolutionary conservation - with visual cues for how each annotation might contribute to pathogenicity. Users can assign a significance to variants, as well as attach freeform text notes. Analyses and variant annotations can be saved and exported, allowing users to return to the analysis at any time. *Gene.iobio* allows a user to perform rapid and comprehensive variant interrogation and prioritization in a single visual and interactive web interface.

We have applied *gene.iobio* in numerous clinical settings at our institution, where it has contributed to the clinical and genetic diagnosis of numerous cases. One representative case was of a boy, less than 10 years old, whose primary objective finding was dysgenesis of the corpus callosum with retrocerebellar fluid collection, laryngeal cleft, cryptorchidism, proximal projections from 2nd metacarpals bilaterally, and non-familial facial profile with ear pits, unusual scalp hair pattern, and abnormal transverse palmar creases. Less specific findings included intellectual disability, autism spectrum disorder, unique pigmentation pattern on torso, and food aversion (Figure 1a). The child and his parents underwent whole exome sequencing through a commercial sequencing provider, who returned a “likely pathogenic” synonymous variant in the report. However, the geneticist following the family disagreed with the report’s findings and requested our group to obtain the raw sequencing data (CRAM files) and perform independent variant calling and variant interpretation.

**Figure 1:**
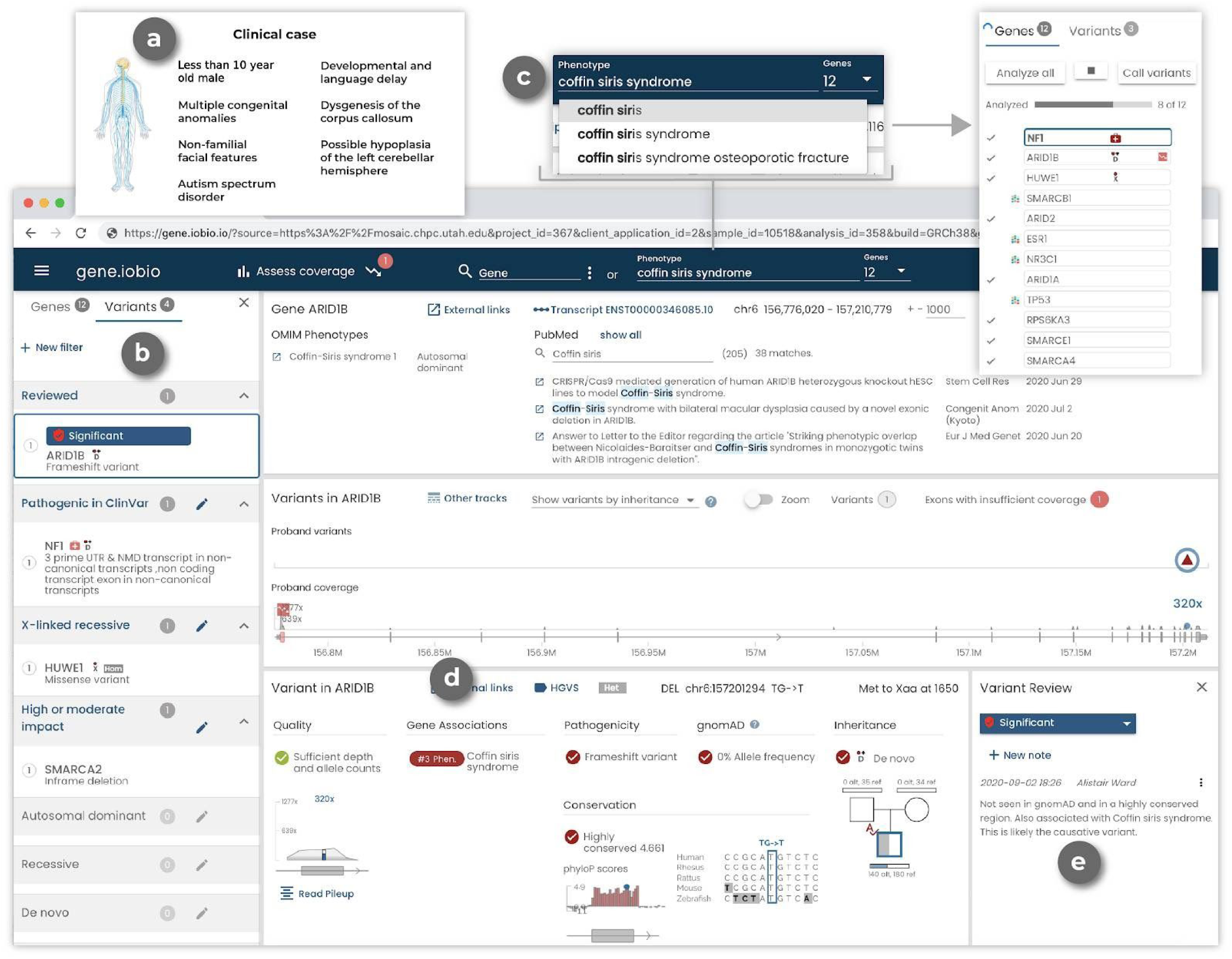
A representative clinical case as viewed in *gene.iobio*. a) Clinical case information and phenotype description (not part of *gene.iobio*). b) Prioritized variants are shown in the left panel and are organized based on their review status, ClinVar pathogenicity, and mode of inheritance. A list of all loaded genes is available in the Genes tab of the left panel. c) Phenotype input components in *gene.iobio* including: generating a list of genes associated with Coffin Siris syndrome (generated by Phenolyzer), OMIM Gene-Phenotype relationships with inheritance mode and a searchable list of PubMed articles associated with the gene. d) Variant details in *gene.iobio* including variant quality, phenotype associations for the current gene (as generated from the phenotype search component), consequence, gnomAD allele frequency, inheritance and nucleotide conservation. e) Variant review capabilities in *gene.iobio* including the ability to assign a significance (Significant, Unknown significance, Not significant, Poor quality, Not reviewed) as well as enter a free form note.

Following this reprocessing and reanalysis, the bioinformatics and clinical genetics teams reviewed candidate variants in *gene.iobio*. One of the candidate variants was a rare de novo frameshift variant in ARID1B. This variant was prioritized at the top of the variant list panel in *gene.iobio* (Figure 1b). Coffin Siris syndrome was part of the initial differential diagnosis for this patient, but the patient did not present with classic Coffin-Siris syndrome. With this consideration, the clinical team entered Coffin Siris syndrome into the phenotype entry component of *gene.iobio*, which uses Phenolyzer^23^ to generate a list of phenotype-associated genes (Figure 1c). *ARID1B* was among the genes in this Coffin Siris syndrome-associated gene list (Figure 1d). Also within this view of *gene.iobio*, the clinical team reviewed key variant annotations such as consequence, gnomAD allele frequency and inheritance (Figure 1d). The clinical team also reviewed OMIM phenotypes and PubMed publications related to *ARID1B* for further clinical evidence. After reviewing this variant, phenotype and literature evidence, the clinical and bioinformatics teams were able to conclude this variant was causative of the patient’s phenotype, as it fits within a larger group of ARID1B-related disorders, of which Coffin-Siris is within^40^. Lastly, the team assigned a significance to the *ARID1B* variant and entered a note describing why this variant was considered causative (Figure 1e).

*Gene.iobio* uses a sophisticated software architecture (see Methods) to provide real-time, comprehensive and visually-driven variant annotations and gene information (Figure 2). *Gene.iobio* takes in a number of inputs, including variant and sequence alignment files (Figure 2a). These genomic data files can be provided from a user’s local machine or as a publicly accessible URL. Files are never directly uploaded or stored, but rather data from these large files are analyzed in a gene region-specific manner, with small discrete data chunks being streamed to backend bioinformatics services (Figure 2b). This allows for real-time analysis and visualization of the genomic data. During an analysis, *gene.iobio* automatically passes numerous pieces of information to various knowledge sources and bioinformatics services, which return pertinent information for variant filtering, interpretation and prioritization (Figure 2c). This robust and versatile approach allows users to iteratively analyze variants, entering new genes and prioritizing new variants. Analyses and variants can be saved and exported for other downstream uses or for returning to *gene.iobio* at a later time.

**Figure 2:**
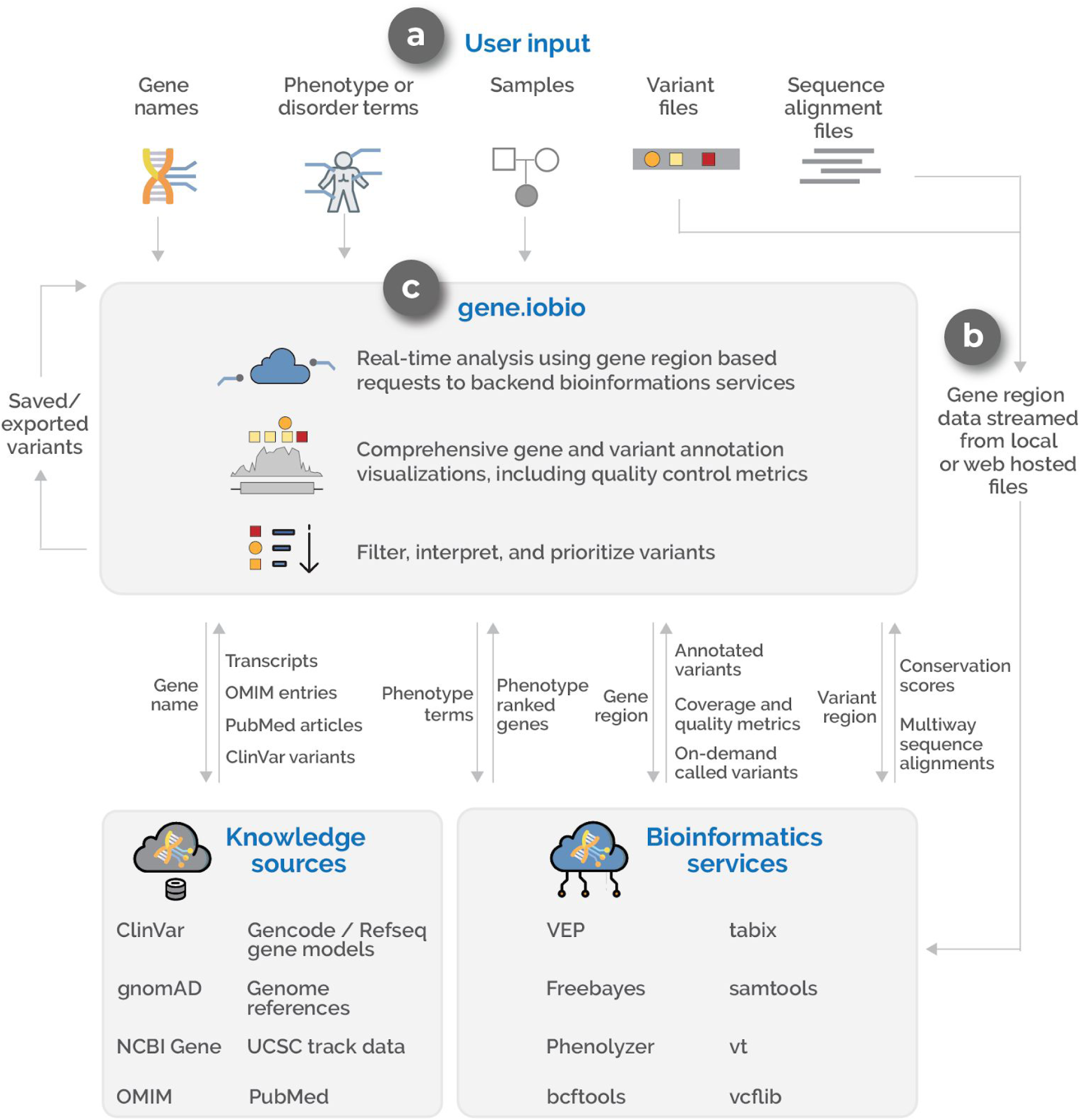
An overview of the *gene.iobio* system and software architecture. a) Inputs for *gene.iobio* include gene names (single or multiple), phenotypes or disorder terms, samples and relatedness between samples, variant files (VCF) and sequence alignment files (BAM or CRAM). b) Gene region data (gene genomic coordinates +/- 1000bp) is streamed from files provided on the user’s local machine or from publicly accessible URLs to a series of backend bioinformatics services. c) *gene.iobio* coordinates information exchanges between knowledge sources (through APIs where available or from custom-built iobio backend services) and bioinformatics services to display variant and gene annotations and draw visualizations.

Clinical expertise informs candidate variant prioritization. This can be especially useful in research settings where clinicians can often be more inclusive in their consideration of candidate variants and is distinct from commercial sequencing providers who typically only have access to a small set of phenotype terms and are more restricted in the variants they report. Clinicians in a research setting can also consider more subtle phenotypes and/or phenotypes that may not have been noted in the initial evaluation. For example, expert clinicians at our institution have reviewed variants in genes prioritized by our computational pipelines and provided key clinical insights about gene:disease association or first hand experience of patients with similar genetic and phenotypic findings. This has included a case of a male between 30 and 40 years old with adult onset leukodystrophy. The clinical team prioritized a rare X-linked recessive missense variant in *ATP6AP2* due to the gene’s association with epilepsy syndromes, an insight that was not immediately available to the bioinformatician analyzing the case (Figure 3a). *Gene.iobio* provided this clinician with a comprehensive and easy to understand summary of the variant as well as the OMIM phenotypes and PubMed literature (Figure 3a). While this particular patient did not display all of the phenotypes associated with X-linked Parkinsonism with spasticity, as defined by OMIM, the patient had sufficient phenotypic overlap with the described disorder that the variant was considered diagnostic. This example highlights the clinical knowledge bases that *gene.iobio* provides to help clinical experts bolster evidence for a given variant’s pathogenicity.

**Figure 3:**
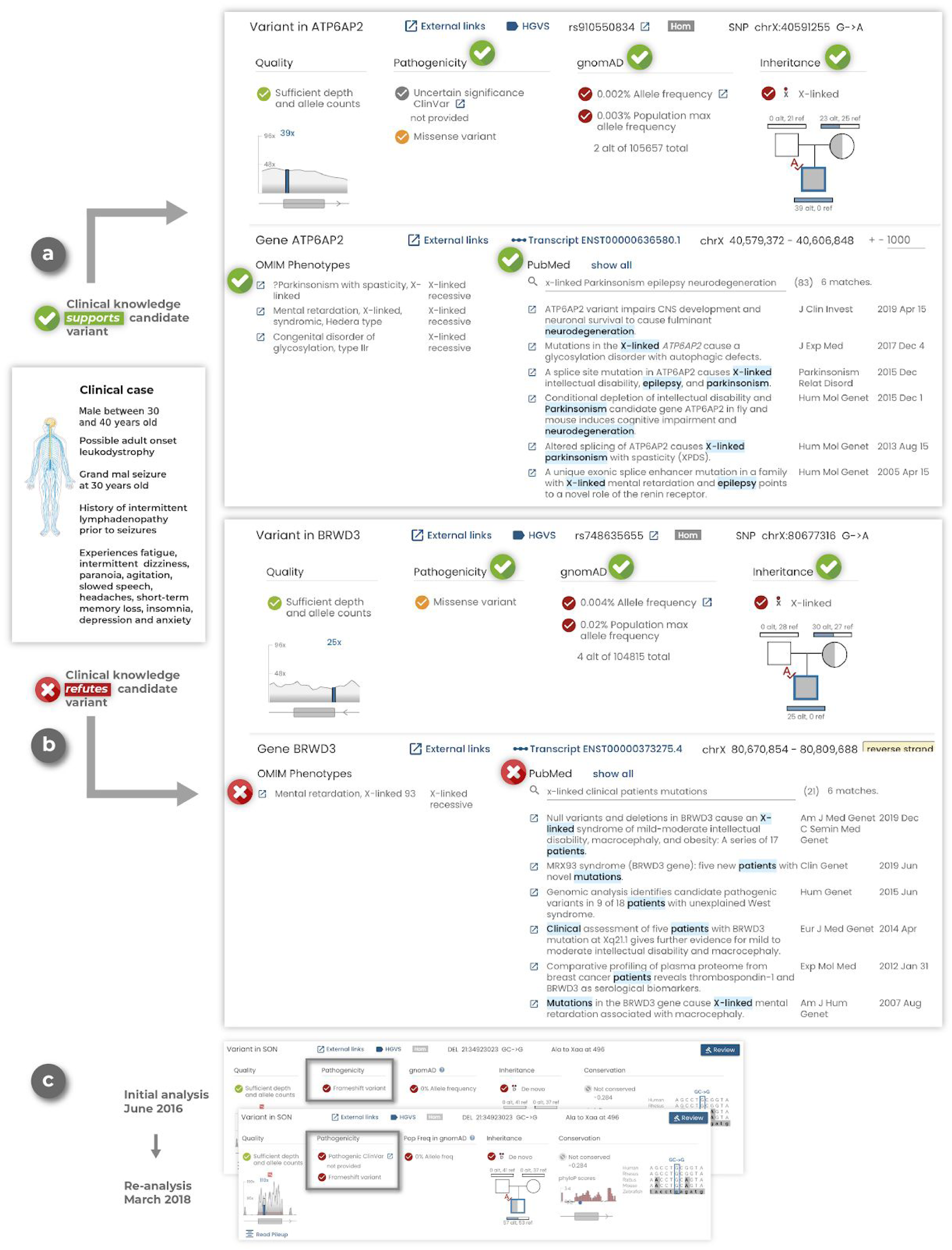
*Gene.iobio* empowers clinical experts during variant prioritization and is a platform for variant reinterpretation. a) In a representative clinical case of a male between 30 and 40 years old with adult onset leukodystrophy, *gene.iobio* provides comprehensive variant and clinical information that supports the candidate ATP6AP2 variant. b) In the same clinical case, *gene.iobio* provides comprehensive variant and clinical information to refute the candidate BRWD3 variant. c) Reanalysis of a previously undiagnosed case in *gene.iobio* reveals an updated pathogenic ClinVar assertion for a candidate variant in the SON gene.

Conversely, clinical experts can also use *gene.iobio* to add evidence against a given candidate variant. In the same case as above, clinical experts used *gene.iobio* to refute a computationally prioritized rare X-linked recessive missense variant in *BRWD3* (Figure 3b). Using *gene.iobio* the clinical team could confirm that while this variant was rare and possibly impactful, the gene is associated with X-linked mental retardation in OMIM and numerous publications in PubMed support this association (Figure 3b). However, this phenotype is not consistent with the patient, who has no described intellectual disability, allowing the clinical team to correctly discard this variant from their analysis. Both previous examples highlight the utility of bringing clinical expertise closer to genomic analyses and variant prioritization, as those responsible for the patient’s clinical care have unique insights about the patient’s phenotypes and specific disease presentation.

*Gene.iobio* is uniquely positioned for reanalysis and reinterpretation of sequencing data and the increasing efforts to improve diagnostic yield. *Gene.iobio* provides users with the most up-to-date variant annotations and gene:disease associations, served from their latest releases through automatically updating backend services (Figure 2c). This allows a user to return to previous sequencing data and analyze variants with the most up-to-date annotations. We have diagnosed numerous cases at our institution using *gene.iobio* to reinterpret variants from previously undiagnosed cases. One example was a case that was sequenced by a commercial provider and remained undiagnosed for multiple years. After obtaining the sequencing data from the commercial provider, the clinical team reviewed computationally prioritized variants in *gene.iobio*. During this review, the clinical team identified a rare de novo frameshift variant in the *SON* gene. Since the original sequencing was performed, numerous publications have described de novo loss of function variants in the *SON* gene^41–44^, with the published patients’ phenotypes largely overlapping that of our patient. Furthermore, the variant had also been asserted as pathogenic in ClinVar. All of this evidence, as displayed in *gene.iobio*, was sufficient to return a genetic diagnosis to the patient and family. This example highlights the ease and power of *gene.iobio* as a variant reanalysis and reinterpretation platform.

*Gene.iobio* also provides users with a clearer understanding of nuanced, and sometimes confusing genomic information. *Gene.iobio* allows users to quickly and easily identify and adjudicate potential false positive and false negative variant calls, a particularly challenging task for de novo variants (Figure 4). Allele balance (the number and ratio of reference and alternate allele observations) can often help adjudicate de novo variants. *Gene.iobio* displays allele balance information in an easily understandable and visual format. Allele balance information in *gene.iobio* was used to identify a false negative de novo variant in the *GRASP* gene, where upstream prioritization tools discarded this variant due to low coverage. Yet upon viewing in *gene.iobio*, the user can readily determine that while the total depth is low (16X), the allele balance is consistent with a heterozygous de novo variant (6 alternate observations and 10 reference observations) and no alternate observations are observed in either parent (Figure 4a). Furthermore, the user can launch the integrated IGV viewer and confirm at the read level that this is a high quality variant and should likely be retained.

**Figure 4:**
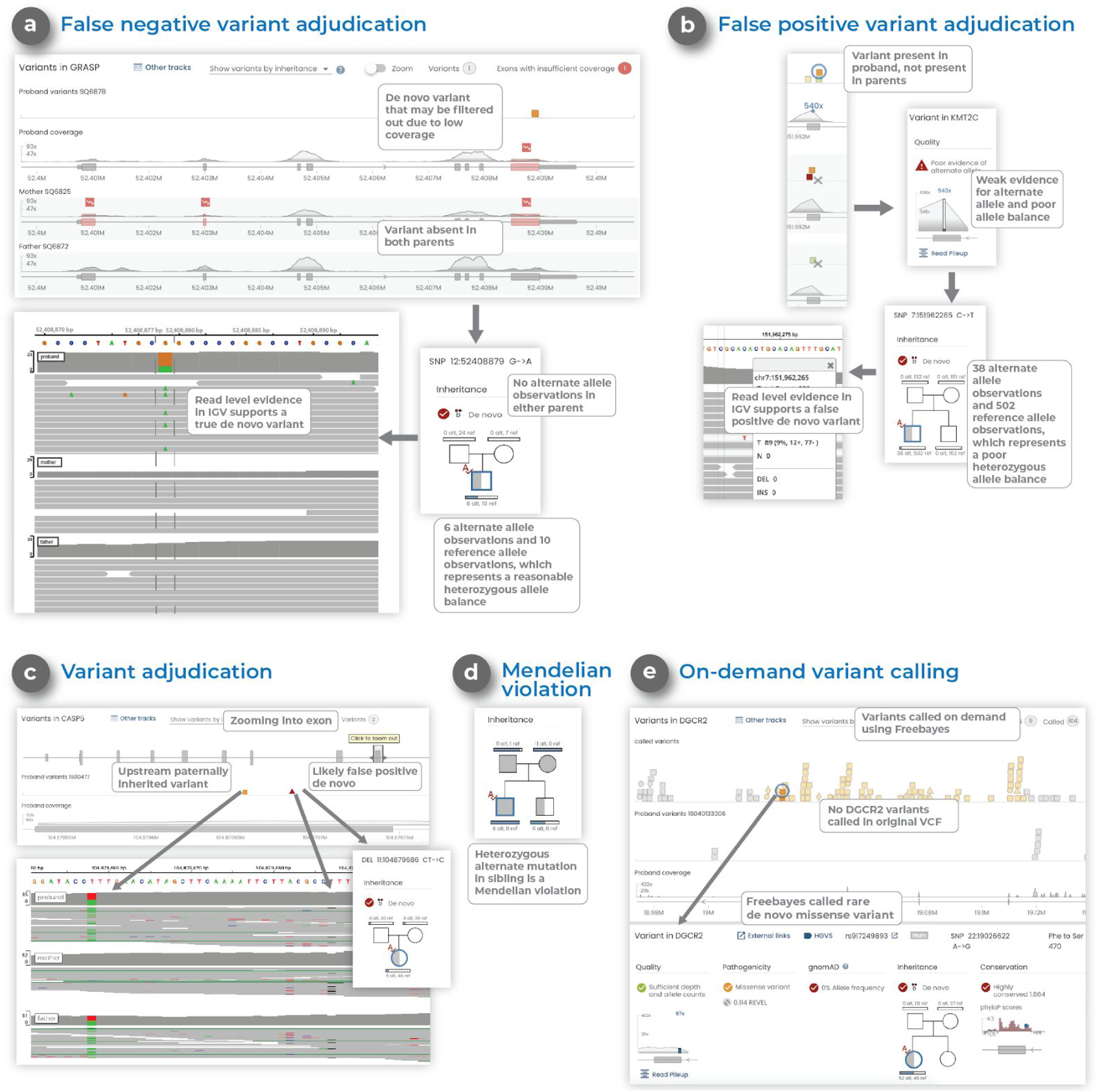
*Gene.iobio* helps adjudicate de novo variants, identify Mendelian violations and provides on-demand variant calling. a) Adjudication of a false negative de novo variant in *gene.iobio* where reasonable allele balance, despite low coverage, suggest the variant could be real. b) Adjudication of a false positive de novo variant in *gene.iobio* where poor allele balance suggests the variant is likely not real. c) Adjudication of a false positive de novo variant in *gene.iobio* where variant evidence is observed in both parents, suggesting the variant could be inherited and not a real de novo. d) A Mendelian violation viewed in *gene.iobio* where a sibling has a heterozygous genotype, despite both parents being homozygous alternate, a clear Mendelian violation. e) On-demand variant calling in *gene.iobio* using Freebayes where a previous variant calling method failed to call any variants in the *DGCR2* gene, but Freebayes variant calling in *gene.iobio* identifies numerous variants within the gene.

Similarly, allele balance views in *gene.iobio* can be used to discard likely false positive de novo variants. One such example was a variant observed in the *KMT2C* gene (Figure 4b). This variant was prioritized as de novo by upstream prioritization tools due to it being heterozygous in proband, and both parents being homozygous reference. However, upon viewing the variant in *gene.iobio*, the user can readily see a poor allele balance and poor evidence for the alternate allele (38 alternate observations and 502 reference observations) in the proband sample. This evidence, provided by *gene.iobio*, suggests this variant is a false positive and should likely be discarded.

Instances also exist where a variant is prioritized as de novo in the proband, yet evidence for the alternate allele exists in the parents, despite their genotypes being reported as homozygous reference. *Gene.iobio* also empowers users to identify these nuanced situations. One such example was observed in the *CASP5* gene, where the proband was genotyped as heterozygous and both parents were genotyped as homozygous reference (Figure 4c). However, when viewing this variant in *gene.iobio*, the user can readily observe that while there are 6 alternate allele observations in the proband, each parent also has 4 alternate allele observations. This evidence can be inspected further in IGV, where a nearby paternally inherited variant can also be observed. This evidence for the alternate allele in both parents and read level inspection in IGV suggests the variant may be inherited from either parent and is likely not a true de novo.

Variant prioritization efforts only consider alleles that segregate with affected status. These Mendelian modes of inheritance include: autosomal dominant; autosomal recessive; de novo and X-linked. Violations of these inheritance modes are clinically important but can also lead to important questions about the samples (if sample swapping has occurred), the underlying genomic region (low complexity regions prone to calling multi-allelic variants) and the variant quality (often due to low complexity or difficult to sequence or call regions). A representative example of a Mendelian violation identified by *gene.iobio* was seen in the *CNGA1* gene (Figure 4d). In this example, the unaffected sibling in a quartet sequencing study shows a heterozygous genotype, despite both parents having strong evidence for their homozygous alternate genotypes. This is a clear violation of Mendelian inheritance modes and would lead the user to question the genotypes of all individuals in the family. This example highlights how *gene.iobio* conveys complex genetic concepts, encoded in genomic files, in a visual format that is immediately intuitive to the user.

In contrast with current variant analysis platforms that rely entirely on the variants called by the variant calling pipeline, *gene.iobio* has built-in variant calling functionality. This approach can be limiting in that variants are often missed or inappropriately removed by post-processing filtering steps. *Gene.iobio* addresses this limitation by directly integrating Freebayes^19^ for on-demand variant calling. This is especially useful for cases where variants are suspected but have not been called, or to provide confirmation that the provided VCF file has not undergone overly restrictive filtering, removing potentially interesting variants. Freebayes variant calling in *gene.iobio* calls variants in the provided gene regions in real-time. One example where this approach was useful was a case where a prior variant calling pipeline failed to call any variants in the gene *DGCR2*, a potentially clinically-relevant gene for the case. However, Freebayes variant calling in *gene.iobio* identified numerous variants (Figure 4e). Our previous publication^45^ also describes the benefits of this variant calling approach in early infantile epileptic encephalopathy cases.

*Gene.iobio* is a comprehensive and feature-rich variant interrogation and prioritization tool that incorporates state-of-the-art bioinformatics tools and clinical genetics resources into a single visual and interactive interface (see Figure S1). This single application approach removes the burden on the user in numerous ways. *Gene.iobio* removes the need to perform complex command line operations and interpret often cryptic bioinformatics file formats and metrics. *Gene.iobio* removes the need to match a given variant to external resources such as publicly reported variants databases, and is publicly-available for academic use (http://gene.iobio.io/) and has been integrated into the Mosaic^46^ tool at our institution.

## Discussion

Genetic information is becoming more routinely used to guide patient care. As such, clinical care providers are taking an increasingly active role in genetic analysis and diagnosis, from independently reviewing genetic findings to performing variant prioritization tasks. However, current variant prioritization and bioinformatics approaches rely almost exclusively on command line tools. Given their training and expertise, it is unreasonable for providers to add command line computational bioinformatics to their current patient care regimen. Our solution to this challenge has been to develop intuitive, visually driven web tools that are immediately usable by clinical care providers. This approach was the motivation for developing *gene.iobio*, a comprehensive genomic analysis and variant review application.

*Gene.iobio* enables secure, clinically-driven variant interrogation and prioritization, bringing clinical care providers’ intimate knowledge of the patient’s disease and phenotype closer to their genetics. *Gene.iobio* is a web application that allows providers to perform variant prioritization tasks through the web browser of any typical computer, without any specialized hardware or software. *Gene.iobio* ensures genomic data security, as data is never uploaded or stored and all data is queried through secure data requests and connections. *Gene.iobio* is also interactive and visually-driven, allowing providers to immediately intuit what the data is representing and directly interact with it, regardless of their bioinformatics experience. Furthermore, visualizations and interactive data are streamed into the application in real-time, allowing users to immediately interact with and analyze their data. This real-time approach analyzes only discrete, user-provided genomic regions, removing the need for large genomics file uploads and for long run time end-to-end data processing. Lastly, *gene.iobio* addresses numerous data insufficiencies issues and many nuanced considerations during variant prioritization. These include the adjudication of putative de novo variants, as well as Mendelian violations - all of which are represented to the user in an easily understandable visual format.

We continue to actively develop *gene.iobio*, releasing regular updates with new features and fixing issues raised by our users. Furthermore, as sequencing and genomic data uses continue to expand, *gene.iobio* is well suited to integrate new features, adopt new annotation metrics and display new data types. Since its initial development, we have added new variant metrics, such as REVEL scores. We anticipate adding pertinent new metrics, such as the metric for constrained coding regions^47^, as they are published. We are also exploring ways to add other relevant functional genomics data types into *gene.iobio*. In the future these may include data for ChIP-seq, RNA-seq ATAC-seq or methylation studies.

*Gene.iobio* reimagines many of the current paradigms in clinical genetics and addresses many of the challenges associated with the increased incorporation of genetic information into clinical care. The level of ease and sophistication provided by *gene.iobio* is unmatched by any other existing tools. We anticipate *gene.iobio* will help lower technical barriers and allow more providers to review and prioritize genetic variants in their own clinical practices and patients. These providers have an intimate understanding of their patient’s disease presentation and phenotypes, information that is unavailable to the bioinformatics analyst in the current paradigm. We anticipate enabling clinicians in this way will help accelerate the adoption of genetic information into clinical care decisions and ultimately improve patient care and outcomes. We also anticipate *gene.iobio* contributing to new genetic diagnoses and the discovery of new genetic disorders through reanalysis and reinterpretation of previously undiagnosed cases, an approach that is being increasingly appreciated in the field.

## Data Availability

https://github.com/iobio/gene.iobio.vue

## Acknowledgements

We would like to acknowledge all those whose feedback, both through personal use and organized testing sessions have been instrumental in focusing *gene.iobio*, and *iobio* development in general. This includes: Rong Mao, Pinar Bayrak-Toydemir, Steven Guthery, Marti Tristani-Firouzi, Nicola Longo, Betsy Ostrander, Hilary Coon, Josh Bonkowsky, Tatiana Tvrdik, Will Dere, Karl Voelkerding, Attila Kumanovics, Karin Chen, Russ Butterfield, Steve Bleyl and David Nix. This research was carried out with funding from NHGRI (R01HG009712 to GTM).

